# Positive and negative stroke signs revisited: dissociations between synergies, weakness, and impaired reaching dexterity

**DOI:** 10.1101/2021.07.21.21260448

**Authors:** Alkis. M. Hadjiosif, Meret Branscheidt, Manuel A. Anaya, Keith D. Runnalls, Jennifer Keller, Amy J. Bastian, Pablo A. Celnik, John W. Krakauer

**Affiliations:** Neurology, Johns Hopkins University, Baltimore, MD; Physical Medicine and Rehabilitation, Johns Hopkins University, Baltimore, MD; Neuroscience, Johns Hopkins University, Baltimore, MD; cereneo Center for Research and Neurorehabilitation, Weggis, Switzerland; Kennedy Krieger Institute, Baltimore, MD; Santa Fe Institute, Santa Fe, NM

**Author notes:** These authors contributed equally to this work (Alkis. M. Hadjiosif, Meret Branscheidt).

## Abstract

Most stroke victims experience motor deficits, usually referred to collectively as hemiparesis. While hemiparesis is one of the most common and clinically recognizable motor abnormalities, it remains under-characterized in terms of its behavioral subcomponents and their interactions. Hemiparesis is comprised of both negative and positive motor signs. Negative signs consist of weakness and loss of motor control (dexterity), whereas positive signs consist of spasticity, abnormal resting posture, and intrusive movement synergies (abnormal muscle co-activations during voluntary movement). How positive and negative signs interact, and whether a common mechanism generates them, remains poorly understood. Here we employed a planar, arm-supported reaching task to assess post-stroke arm dexterity loss, which we compared to the Fugl-Meyer stroke scale; a measure primarily reflecting abnormal synergies. We examined 53 patients with hemiparesis after a first-time ischemic stroke. Reaching kinematics were markedly more impaired in patients with subacute (<3 months) compared to chronic (>6 months) stroke even when matched for Fugl-Meyer score. This suggests a dissociation between abnormal synergies (reflected in the Fugl-Meyer scale) and loss of dexterity, which in turn suggests different underlying mechanisms. Moreover, dynamometry suggested that Fugl-Meyer scores capture weakness as well as abnormal synergies, in line with these two deficits sharing a neural substrate. These findings have two important implications: First, clinical studies that test for efficacy of rehabilitation interventions should specify which component of hemiparesis they are targeting and how they propose to measure it. Second, there may be an opportunity to design rehabilitation interventions to address specific subcomponents of hemiparesis.

## Introduction

Stroke is one of the leading causes of disability globally, with an estimated 13.7 million individuals suffering a stroke each year (Johnson et al. 2019). A large fraction of strokes (up to 70-80%) results in some degree of motor impairment (Nakayama et al. 1994; Rathore et al. 2002; Parker, Wade, and Hewer 1986), which makes activities of daily living harder, compromising quality of life (Niemi M L et al. 1988; Viitanen et al. 1988). Hemiparesis (or upper motor neuron syndrome) is clinically quite recognizable but remains surprisingly under-characterized in terms of its behavioral components. It has been known since the late 19^th^ century that hemiparesis is comprised of loss of ability (negative signs) and an intrusive movement disorder (positive signs) (Hughlings Jackson 1884; Pearce 2004). Negative signs consist of weakness and loss of dexterity or fractionated motor control, whereas positive signs consist of spasticity, abnormal resting postures, and abnormal synergies whereby multiple muscles or joints become co-activated during voluntary movement. How positive and negative signs relate to each other remains poorly understood. Bridging this knowledge gap will be essential for the treatment of hemiparesis, as it will allow us to (a) different components of hemiparesis, allowing clinicians to more reliably track motor recovery after stroke, (b) better isolate and target individual components to make rehabilitation more effective, and (c) better assess the efficacy of rehabilitation interventions.

Among positive signs of stroke, abnormal synergies have been the focus of particular attention and widely recognized as a crucial characteristic of motor impairment after stroke. Thomas Twitchell’s classic work (Twitchell 1951) describes the time course of recovery of voluntary movement after stroke, from plegia to flexor then extensor synergies to out-of-synergy movements. Given that recovery can get stuck at any point along this sequence, Signe Brunnstrom (Brunnstrom 1966) suggested therapeutic procedures to increase the chance of a patient progressing through it. A scale was subsequently developed to measure and track recovery from synergies – the Fugl-Meyer scale (Fugl-Meyer et al. 1975).

The negative signs of stroke are weakness and loss of dexterity or motor control. Dexterity generally refers to the ability to flexibly and independently control muscles and joints to generate the movement repertoire required by a given task. Importantly, the term should not be considered synonymous with or reserved for finger movements. Dexterity might require practice and, after stroke, it can recover independently of weakness (Cortes et al. 2017; Xu et al. 2017). Notably, the intrusion of abnormal synergies might mask dexterity. A previous study showed that when patients with chronic stroke made 3D reaching movements within synergy, trajectories appeared comparable to those made by healthy controls (Zackowski et al. 2004). In contrast, when these same patients attempted an out-of-synergy reach that required elbow extension, the trajectory was degraded by an intrusive flexor synergy. This intrusion is thought to be invoked in part by the patient needing to lift their arm against gravity, as several studies have shown that external support of the weight of the arm can reduce the effect of abnormal synergies and increase the arm’s available workspace (Beer et al. 2007; 2004; Sukal, Ellis, and Dewald 2007). Thus, here we examined the relationship between abnormal synergies and loss of arm dexterity after stroke. To obtain a measure of arm dexterity loss, we quantified kinematics in a planar reaching task. The apparatus provided full support of the weight of the arm, allowing us to assess arm dexterity while minimizing weakness and the intrusion of synergies; moreover, the apparatus constrained the trunk, minimizing the use of compensatory strategies. To measure the extent of abnormal synergies, we used the Fugl-Meyer scale for the upper extremity (FM-UE), which was specifically designed to quantify abnormal synergies post-stroke (Fugl-Meyer et al. 1975; Brunnstrom 1966; Twitchell 1951). Because recovery of reaching dexterity and abnormal synergies may have different time courses, we compared them at two different times post-stroke (Bernhardt et al. 2017): during the early and late sub-acute stage (up to 3 months post-stroke) and the chronic stage (at least 6 months post-stroke). We also assessed weakness in a subset of the patients using dynamometry.

## Materials and Methods

### Participants and Ethics Statement

Participants were recruited as part of a multiple-task study of the motor learning, control, and physiology of stroke patients (PaLaS study, Physiology and Learning after Stroke). The study compared these modalities between two stages in recovery: subacute (< 3 months post-stroke) and chronic (> 6 months post-stroke). Table 1 shows details for each of the 53 stroke patients (27 subacute, 26 chronic) included in this paper, whereas Table 2 shows summary demographics and assessment metrics for the two patient groups and 17 healthy, age-range-matched controls (age comparisons between controls and either patient group, or between patient groups: all p>0.3). Recruitment and data collection took place in Johns Hopkins University and the Kennedy Krieger Institute in Baltimore, MD from December 2015 through February 2020; participant flow through the study is shown in Figure 1. Patients were recruited from the stroke and rehabilitation units at Johns Hopkins, previous study participants, respondents to advertisement (flyers posted within the hospital), and stroke support groups to which the study was advertised. Healthy controls were recruited through advertisement and among previous study participants. All participants received monetary compensation for their time ($20/hour). Participants provided informed consent in accordance with the Declaration of Helsinki, whereas procedures were approved by the Johns Hopkins Institutional Review Board.

**Table 1.** Patient characteristics at T1. Showing time since stroke, lesion side, age (non-overlapping 5-year range), gender, Fugl-Meyer Assessment for the Upper Extremity (FM-UE, max. 66); Montreal Cognitive assessment (MoCA, max. 30), and Action Research Arm Test (ARAT, max. 57). ►

| Time since stroke | Lesion Side | Age (5-y range) | Gender | MoCa | FM-UE | ARAT |
| --- | --- | --- | --- | --- | --- | --- |
| **Subacute patients** |  |  |  |  |  |  |
| *(days)* |  |  |  |  |  |  |
| 29 | R | 31-35 | M | 23 | 59 | 55.5 |
| 8 | L | 61-65 | F | 22 | 53 | 39 |
| 17 | R | 66-70 | F | 23 | 55 | 57 |
| 15 | R | 66-70 | F | 20 | 56 | 45 |
| 12 | R | 66-70 | M | 20 | 59 | 45 |
| 30 | R | 21-25 | F | 27 | 64 | 57 |
| 57 | L | 56-60 | M | 25 | 33.5 | 36 |
| 17 | L | 41-45 | F | 20 | 53 | 34.5 |
| 8 | R | 61-65 | M | 21 | 52.5 | 41 |
| 57 | L | 26-30 | M | 28 | 61.5 | 57 |
| 19 | L | 36-40 | F | 26 | 58 | 54 |
| 47 | R | 51-55 | M | 24 | 57 | 56 |
| 9 | L | 61-65 | M | 19 | 56 | 48.5 |
| 12 | R | 81-85 | F | 22 | 38 | 37.5 |
| 43 | R | 71-75 | F | 27 | 21 | 9 |
| 47 | R | 61-65 | F | 30 | 63.5 | 57 |
| 6 | L | 61-65 | M | 20 | 65 | 57 |
| 55 | R | 51-55 | M | 24 | 44 | 37 |
| 21 | L | 51-55 | F | 22 | 64.5 | 57 |
| 19 | R | 91-95 | M | 27 | 26 | n.d. |
| 70 | R | 66-70 | F | 28 | 59.5 | 56 |
| 47 | R | 71-75 | F | 23 | 45 | 54 |
| 14 | L | 61-65 | F | 25 | 61 | 55.5 |
| 17 | R | 46-50 | M | 30 | 62.5 | 57 |
| 12 | L | 71-75 | F | 25 | 57.5 | 56.5 |
| 58 | L | 46-50 | M | 29 | 15 | 11 |
| 51 | R | 56-60 | M | 27 | 11.5 | 3 |
| **Chronic patients** |  |  |  |  |  |  |
| *(months)* |  |  |  |  |  |  |
| 27 | R | 26-30 | F | 30 | 58 | 44.5 |
| 54 | R | 41-45 | M | 30 | 13 | 3 |
| 42 | R | 66-70 | M | 27 | 60 | 57 |
| 27 | R | 66-70 | M | 25 | 13 | 6 |
| 27 | L | 56-60 | M | 25 | 58 | 54.5 |
| 39 | R | 56-60 | M | 25 | 47.5 | 44 |
| 24 | R | 46-50 | M | 29 | 12 | 3 |
| 76 | R | 61-65 | M | 28 | 10 | 7 |
| 11 | R | 51-55 | F | 23 | 62 | 57 |
| 88 | R | 61-65 | F | 25 | 17 | 3 |
| 24 | R | 56-60 | M | 20 | 29.5 | 5.5 |
| 8 | R | 76-80 | M | 20 | 50 | 43 |
| 59 | R | 46-50 | M | 22 | 24 | 42 |
| 20 | R | 56-60 | M | 20 | 17.5 | 3 |
| 72 | L | 36-40 | M | 27 | 60.5 | 55 |
| 58 | L | 51-55 | F | 21 | 9 | 2 |
| 35 | R | 51-55 | M | 26 | 64.5 | 55.5 |
| 7 | L | 66-70 | M | 28 | 64.5 | 57 |
| 7 | R | 46-50 | M | 29 | 27 | 14 |
| 8 | R | 51-55 | F | 28 | 36 | 14.5 |
| 17 | R | 56-60 | F | 26 | 13.5 | n.d. |
| 9 | R | 41-45 | F | 28 | 33 | 25.5 |
| 7 | L | 36-40 | M | 26 | 61 | 57 |
| 59 | R | 56-60 | F | 23 | 18.5 | 3 |
| 95 | L | 66-70 | F | 27 | 64 | 57 |
| 103 | L | 36-40 | F | 27 | 48.5 | 52.5 |

**Table 2.** Demographics and clinical characteristics of participants. ± indicates standard deviation across participants. FM-UE: Fugl-Meyer Assessment for the Upper Extremity; ARAT: Action Research Arm Test; MoCA: Montreal Cognitive assessment.

|  | Subacute stroke patients | Chronic stroke patients | Controls |
| --- | --- | --- | --- |
| **N** | 27 | 26 | 17 |
| **Age** | 58.8±16.3 | 54.6±11.5 | 57.6±12.4 |
| **Gender** | 13M/14F | 16M/10F | 8M/9F |
| **Affected Side** | 11R/16L | 7R/19L | n/a |
| **Handedness** | 24R/1L/2A | 24R/2L | 15R/1L/1A |
| **FM-UE** | 50.1±15.6 | 37.4±21.1 | 66±0 |
| **ARAT** | 45.1±15.9 | 30.6 ±23.6 | 57±0 |
| **MoCA** | 24.3±3.3 | 25.6±3.1 | 27.9±1.6 |
| **Time Since Stroke** | 29.5±19.8 days | 38.6±29.6 months | n/a |

**Figure 1:**
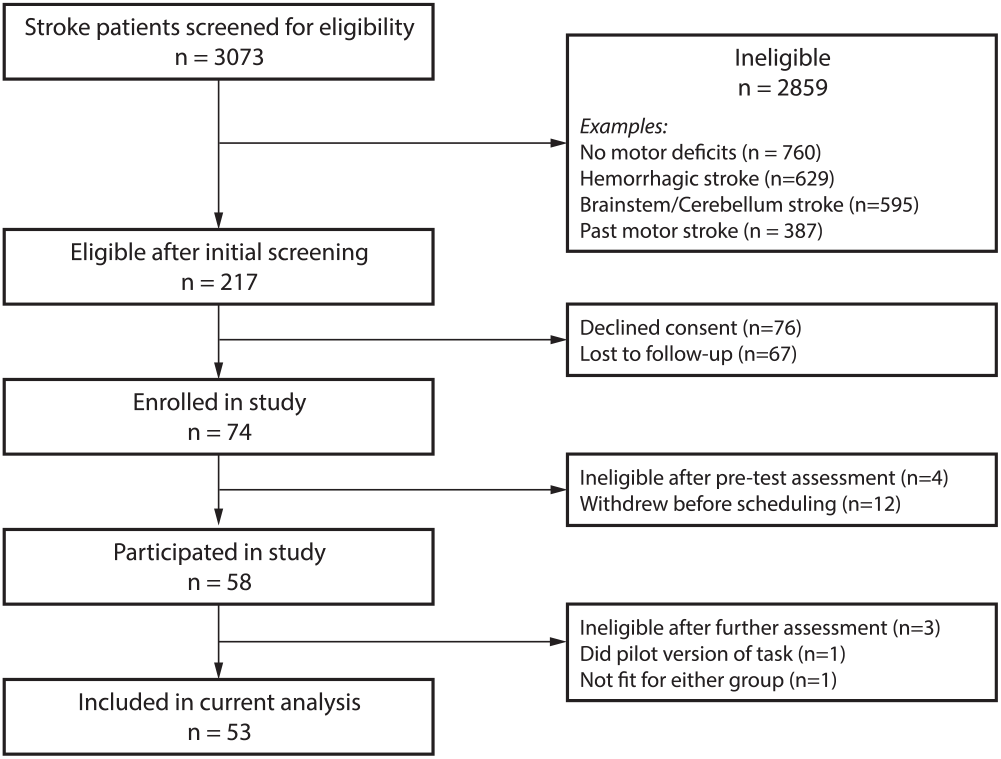
Participant flow through the study. Note that the same patient might fulfill more than one ineligibility criteria.

### Eligibility criteria

Patients recruited had to be over 21 years old, had suffered an ischemic supratentorial stroke that was their first stroke with motor deficits, exhibited some movement with the affected arm, and be able to provide informed consent and understand the tasks involved. Exclusion criteria relevant to the tasks described in this paper included hemorrhagic transformation or associated intracranial hemorrhage; severe congestive heart failure; unstable angina; uncontrolled hypertension; dementia (assessed based on the Montreal Cognitive Assessment, MoCA (Nasreddine et al. 2005)); severe aphasia or ideomotor apraxia, neglect or hemianopia; and orthopedic or pain issues.

### Sessions

Participants underwent two sessions: the main session (T1) and a one-month follow-up (T2). For the subacute group, T1 took place 29.5±19.8 days post-stroke (average±standard deviation) and T2 took place 39.5±8.5 days later. For the chronic group, T1 took place 38.6±29.6 months post-stroke and T2 took place 39.2±7.8 days later.

### Impairment assessment using the Fugl-Meyer scale for the upper extremity (FM-UE)

Assessments were separately scored by at least two different raters (J. Keller, AMH, MB). To obtain the final value, scores were averaged between reviewers (hence some having decimal values). In cases of substantial score differences (3 points or more), assessment videos were again reviewed by both raters together. For the analysis comparing changes in arm dexterity and FM-UE from T1 to T2 (Figure 6), a participant was excluded from the chronic group due to missing FM-UE score. We used the entirety of the score (0-66) for our main analysis. At certain points, as mentioned in our results, we additionally performed comparisons based on subcomponents of FM-UE scores focusing on (a) items referring to movement of the proximal arm (items 3-17 and 31-33 – i.e. everything apart from parts I/VI (reflexes), VII (wrist), and VIII (hand)), (b) items related to within-synergy movements (part II of FM-UE, “Flexor Synergy” items 3-8), or (c) items related to out-of-synergy movements (parts IV and V of the FM-UE: “Movement combining synergies” and “Movement out of synergy”).

### Lesion location

A large fraction of our participants (34 out of 53) had clinical MRI images available, which enabled us to compare lesion size between subacute and chronic populations. Lesion size was significantly larger in the chronic participants for which images were available compared to subacute patients (56794±21612 voxels for chronic (N = 13) vs. 8613±3072 voxels for subacute (N = 21), p = 0.047), in line with lower FM-UE scores in this group (Table 2). Interestingly, in subacute and chronic patients with mild/moderate FM-UE scores (FM-UE≥26, on which our main behavioral analysis focused), two populations with relatively matched FM-UE scores (46.3±4.4 for the 10 chronic vs. 54.4±2.5 for the 19 subacute patients with available imaging, p = 0.17) lesion size was still (marginally) larger in the chronic group (61029±26714 voxels for chronic (N = 10) vs. 9269±3364 voxels for subacute (N = 19), p = 0.086), as shown in Figure 2.

**Figure 2:**
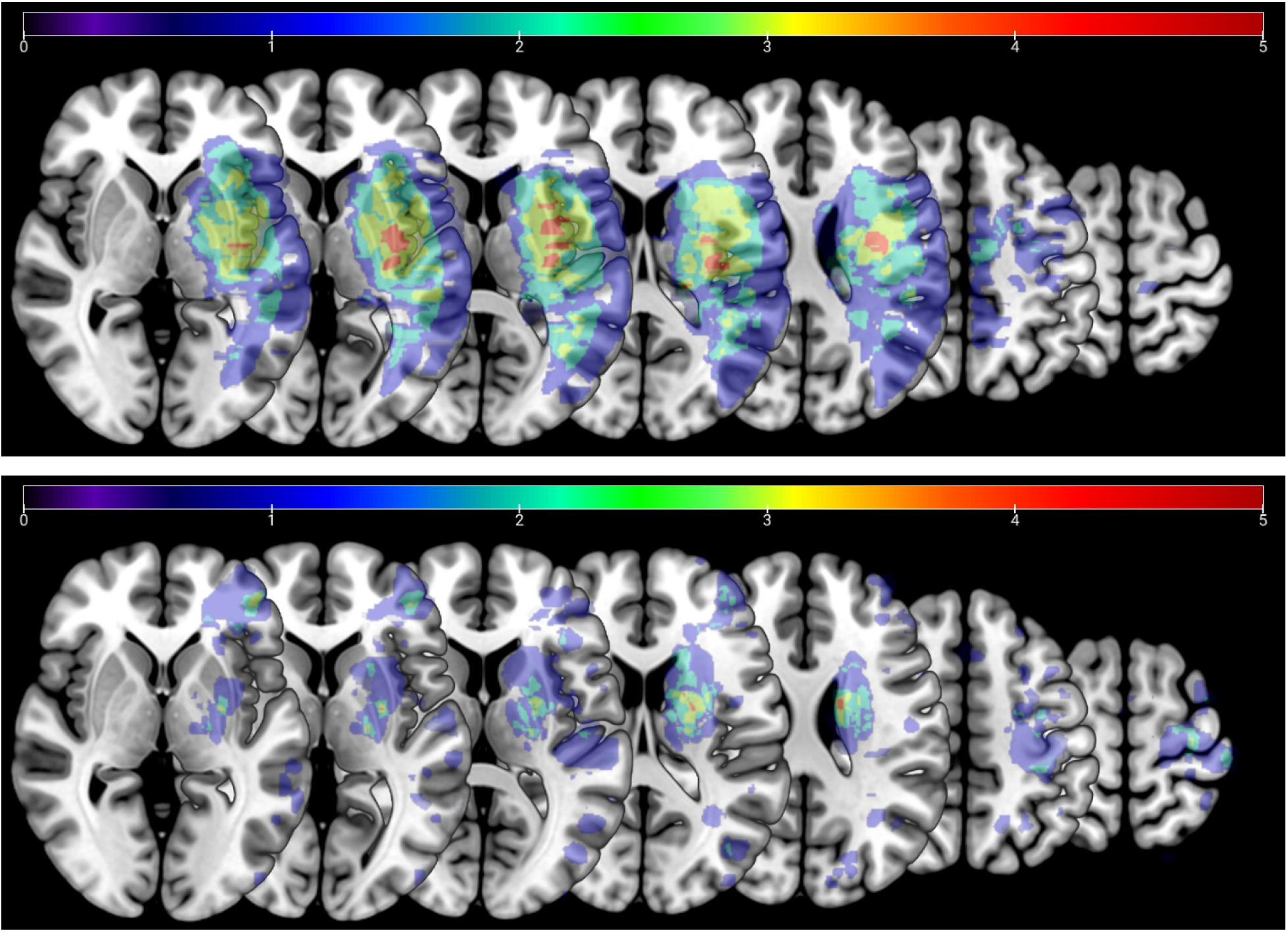
Lesion distribution overlay for chronic (top, N=10) and subacute (bottom, N=19) patients with moderate/mild FM-UE impairment (FM-UE ≥26). Averaged lesion distribution mapped to JHU-MNI space, with lesion flipped to one hemisphere. Color bar indicates patient count.

### Reaching Task Details

Participants sat on a robotic chair (Kinarm exoskeleton), which provided arm support while allowing for planar motion (Figure 3). The chair was positioned against a screen that occluded vision of the arm but allowed projection of targets, and a cursor indicating hand position, at arm level. Participants made 10-cm reaching point-to-point movements to eight targets arranged at 45° intervals about a start position as shown in Figure 3B. Targets and start position were 1cm in radius, where the cursor was 0.5cm in radius. The start position was defined relative to each participants’ shoulder midpoint, and was typically (in 47 out of 53 participants) 45 cm from it but could range from 45-50cm to accommodate different sizes and positioning of participants. Upon positioning the cursor on the start position, a cyan target would appear in one of the eight different target positions. Participants were instructed to initiate a movement to the target soon after it appeared. A movement timer would begin as soon as the participant had moved outside the start position.

**Figure 3.**
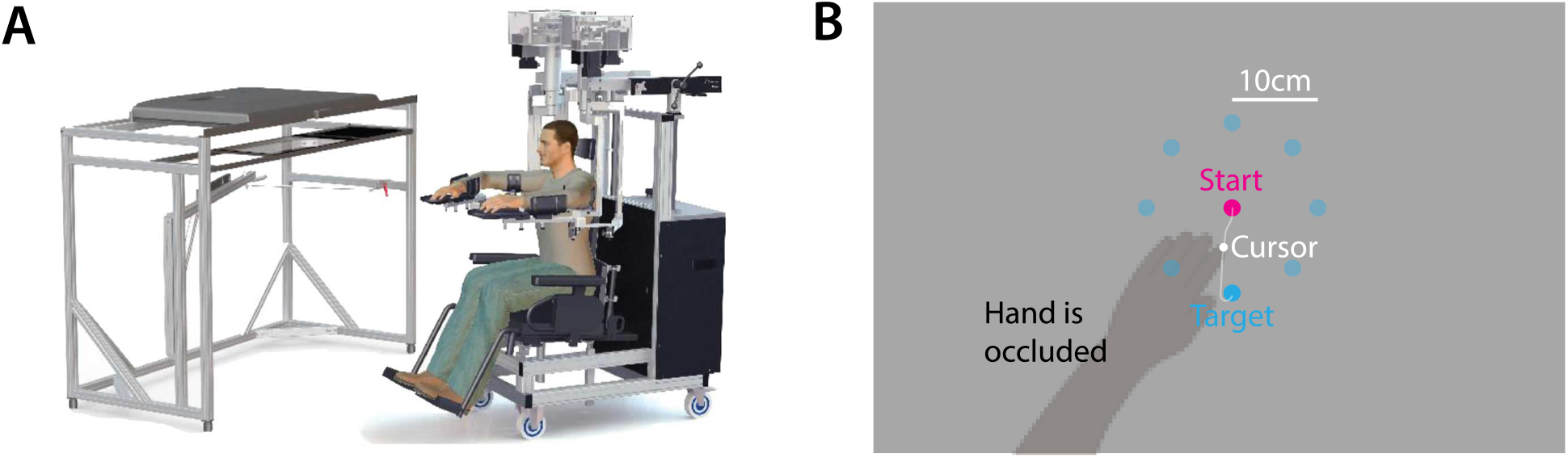
Task design and example trajectories.: Illustration of the experimental apparatus (Kinarm, BKIN Technologies, Kingston, Ontario, figure adapted from (Tyryshkin et al. 2014). **B:** The task: participants had to make point-to-point outwards reaching movements to 8 different targets (cyan) beginning from a starting position (pink). Vision of the hand was occluded; instead, during movement, a cursor was displayed on screen on the plane of the arm.

The movement would end either when the participant reached the target, or an 800ms timer ran out. Upon movement end, the cursor would freeze momentarily and the target would change color based on the participant’s speed. Specifically, it would change to orange if the movement terminated on the target but too quickly (movement time <200ms), or red if the movement failed to reach the target before the 800ms timer ran out. If the cursor reached the target within the desired 200ms to 800ms from onset, and was held inside the target for an additional 500ms, the target would turn green. After an additional 250ms wait time, Kinarm would actively return the arm back to the starting position.

Participants completed four blocks of 88 movements each (11 to each of the eight different targets), beginning with two blocks with the paretic arm, followed by two blocks with the non-paretic arm. Breaks were given between the blocks as necessary. With the exception of one participant, who did the paretic and non-paretic measurements on different days due to scheduling constraints, the entirety of each session took place on the same day and typically lasted about 45 minutes.

### Dynamometry

We measured strength by dynamometry in 25 of our participants (not all of them as it was only later added to the study assessments). Participants sat on a chair, with their trunk straight and their forearm supported on a table in pronation with the elbow at 90 degrees and the shoulder in an open packed position (approximately 60 degrees of abduction and 30 degrees of horizontal adduction) (Bohannon 1990). Using a handheld dynamometer (MicroFet 2, Hoggan Scientific), we measured the maximum effort of four muscle groups: elbow flexors, elbow extensors, shoulder (horizontal) adductors, and shoulder (horizontal) abductors. The average of two trials were taken for each condition. Both the paretic and non-paretic arms were tested, and strength was expressed as a % of the force exerted with the non-paretic arm.

### Data Analysis

Analysis was performed using Matlab (Mathworks, Natick MA). Position data were smoothed using an 8^th^-order, 8 Hz low-pass Butterworth filter, and differentiated to obtain velocity. For the purpose of analysis, we estimated movement onset using a method similar to what described previously in (Cortes et al. 2017): we identified the time of peak speed (first zero-crossing of acceleration that is >8cm/s), and then, going backwards, identified movement onset as the time speed surpassed 2 cm/s. We identified movement end by going forward from the time of peak speed and finding the moment when speed remained <2cm/s for more than 0.1s.

#### Data exclusion criteria

On occasion, after setting up a participant on the robotic chair, reaching some of the targets was impossible because of mechanical constraints. We thus excluded these targets from our analysis. This case was rare: targets were excluded in only two out of 53 participants. From the remaining trajectories, we excluded movements in which (i) movement direction at peak speed was ≥90° away from target direction, (ii) the participant had not moved beyond 30% of the target distance (the criteria used in (Cortes et al. 2017) and previously (Kitago et al. 2015)), or (iii) the movement onset analysis described in the previous paragraph failed to provide an estimate of movement onset. For the second training block that we focus on in this manuscript, these criteria excluded a further 3.72% of patients’ movements on the paretic side and 1.30% on the non-paretic side (for healthy controls, the corresponding value was 0.07% for across both sides).

#### Functional Principal Component Analysis

To assess the quality of kinematics, we used functional principal component analysis (fPCA), a method which applies principal component analysis to functional data (Goldsmith and Kitago 2016). This data-driven analysis allows trajectories to be evaluated without prior assumptions about which trajectory features ought to be emphasized. Here, we used fPCA to estimate the Mahalanobis distance of patients’ trajectories from the reference population of healthy, age-matched controls. For each patient, we averaged these distances into the average squared Mahalanobis distance (AMD^2^) for sessions T1 and T2. Details about this analysis have been reported previously (Goldsmith and Kitago 2016).

#### Estimation of baseline value for AMD^2^ in controls

The functional principal component analysis described above compares patients’ trajectories to the population of corresponding trajectories from healthy controls. Because of variability within the reference population itself, AMD^2^ scores would be nonzero for controls themselves. To estimate an AMD^2^ value for this baseline, we calculated AMD^2^ between the trajectories of each control participant compared and the trajectories of all other controls.

#### Statistical comparisons

To compare AMD^2^ between subacute and chronic patients we used unpaired, 2-tailed t-tests without assuming equal variances (Welch’s t-test using the Satterthwaite approximation for effective degrees of freedom); p-values reported in Results were based on these tests. In a secondary, non-parametric analysis, we compared these two groups also using a bootstrap procedure (Efron and Tibshirani 1994) using 10,000 permutations within each subgroup, which also yielded significant differences between subacute and chronic patients’ AMD^2^.

For comparisons within the non-paretic data, we first used an ANOVA to investigate any effect of group (subacute, chronic, controls) since there was no prior expectation either patient group would show reduced reaching dexterity (higher AMD^2^) in the first place. After the ANOVA, we used a Tukey-Kramer test for multiple comparisons for post-hoc tests.

## Results

### Subacute stroke patients had worse reaching dexterity compared to chronic stroke patients despite matched Fugl-Meyer scores

To measure quality of reaching movements, we had participants perform 10cm point-to-point reaching movements to eight different targets on the 2D plane with arm support, using the Kinarm Exoskeleton (Figure 3). Online visual feedback was provided in the form of a cursor and, with the help of color cues, participants were prompted to reach and stop at the target within 200-800ms after movement onset (for details, see Materials and Methods). In each session, a total of 176 movements were performed with each hand (two blocks of 11 reaches to each of the eight targets, with paretic-arm blocks performed first).

To avoid familiarization effects (see Figure S1), we focused our analysis on the second block (88 movements). Examples of participants’ trajectories with the paretic arm are shown in Figure 4A. We made two primary observations. First, subacute participants had markedly worse trajectories compared to chronic participants even when matched for FM-UE scores. Second, there was convergence onto the shape of trajectories of the control population as patients’ FM-UE scores improved.

**Figure 4.**
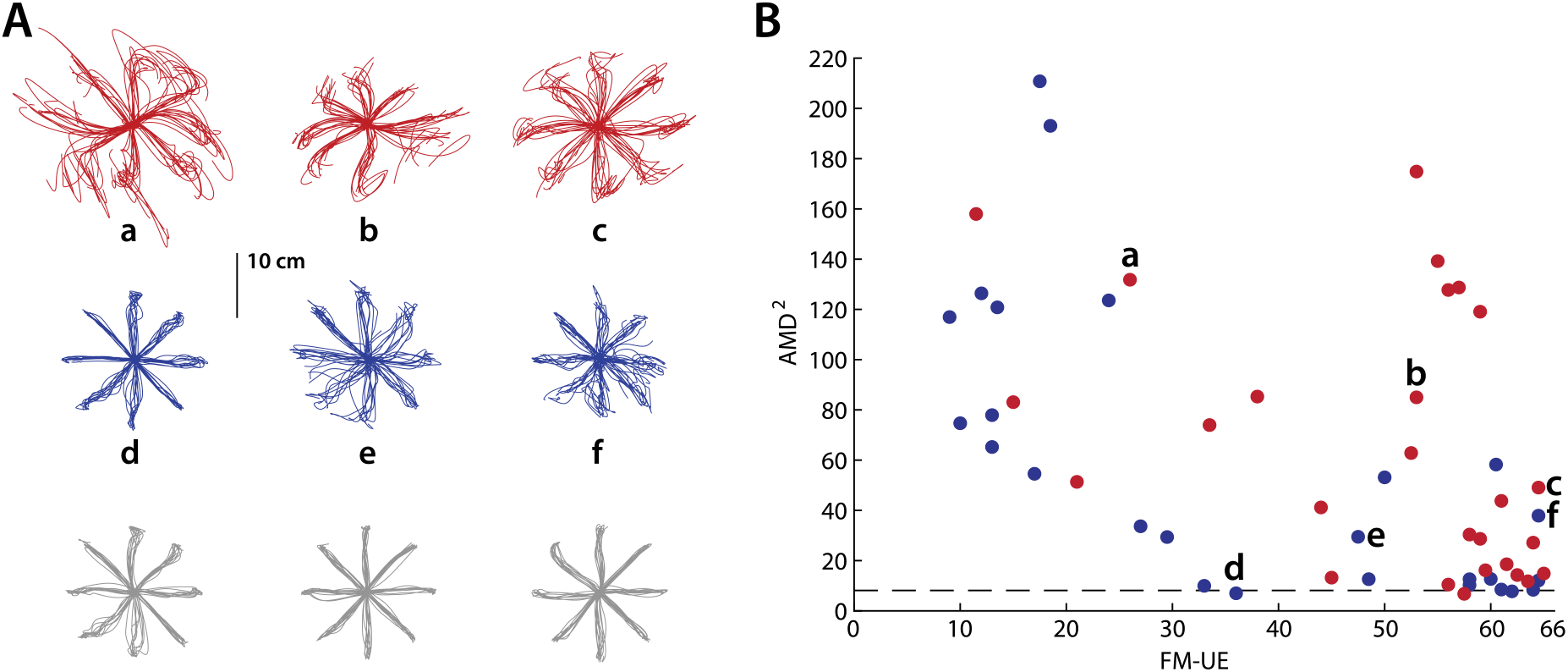
Subacute patients had worse kinematics compared to chronic patients for similar Fugl-Meyer scores. **A:** Exemplar movement trajectories, using the paretic arm, for three subacute patients (red), three chronic patients (blue), and three controls (gray). **B:** Scatter plot of kinematic abnormality (AMD^2^) vs. FM-UE for the subacute (**red**) and chronic (**blue**) groups. Note the higher AMD^2^ for subacute patients, especially the ones with moderately and mildly impaired FM-UE (≥26). The lowercase letters (a-f) point to the corresponding trajectories on the panel **A.** The black dashed line indicates baseline calculated based on control data.

To formally assess reaching trajectories in stroke patients and generate a scalar comparison metric, we used functional principal component analysis (fPCA), a method which compares patients’ movement trajectories to those of a reference population (Kitago et al. 2015; Goldsmith and Kitago 2016; Cortes et al. 2017). In this study, our reference population consisted of a group of 17 age-matched controls (see Table 2). Specifically, fPCA assigns a score to each trajectory produced by the patient, based on how that trajectory’s shape differs from the average control trajectory, given the natural variability of control trajectories (for more details see Materials and Methods). This resulting score is a Mahalanobis distance (MD), and can be understood as a generalization of the z-score; it is large when the movement is impaired (i.e. it is dissimilar to the healthy controls used as a reference) and small when the movement is similar to healthy controls. This analysis avoids having to assess many different kinematic variables (e.g. angular error, accuracy, jerk, curvature), and then not knowing how to interpret them when they dissociate (Kitago et al. 2015; Krakauer and Carmichael 2017). As a measure of reaching dexterity for each participant, we calculated these MDs across trials and target directions. We refer to the resultant measure as the *Average Squared Mahalanobis distance* (AMD^2^) (Cortes et al. 2017).

Because AMD^2^ estimates the dissimilarity of each trajectory from the average trajectory in the reference population, it will be non-zero even for unimpaired trajectories given the natural variability of movement. Hence, we estimated a baseline value for AMD^2^, denoted by how low the AMD^2^ score would be for unimpaired performance, by taking each control participant’s trajectories and computing their AMD^2^ against the trajectories of remaining controls, which yielded an average AMD^2^ of 8.13±0.58 (mean±SEM across control participants).

Consistent with the trajectory shapes shown in Figure 4A, AMD^2^ scores indicated impaired trajectory kinematics for stroke patients compared to controls (average±SEM AMD^2^ for all patients: 61.4±7.5; for all controls: 8.13±0.58). Interestingly, however, subacute patients tended to show markedly worse kinematics than chronic patients despite similar or higher FM-UE scores, as illustrated in Figure 4B. Because very few participants in the subacute stage had low FM-UE scores, resulting in a higher average FM-UE for the subacute group compared to the chronic group (on average, FM-UE of 50.1±3.0 for the subacute vs. 37.4±4.1 for the chronic group, see Figure 4B), we performed additional analysis on participants with moderate and mild impairment (FM-UE≥26, the cut-off is based on previous work (P. W. Duncan, Lai, and Keighley 2000; Krakauer and Carmichael 2017)). For these patients (24 subacute and 16 chronic), despite matched FM-UE (54.3±2.1 for the subacute vs. 51.5±3.3, mean±SEM for the chronic patients, respectively, p = 0.47), trajectory abnormalities were substantially greater in the subacute group (AMD^2^ of 60.6±10.4 vs. 21.5±4.2, p = 0.0016, Figure 5). These two groups were significantly different (p=0.0028) even when the subacute participant with the highest AMD^2^ (174.9, Figure 5, right) was excluded as potential outlier; they were also clearly different in a secondary, non-parametric analysis using bootstrap (Figure S2). We also considered whether this AMD^2^ difference between subacute and chronic patients could be due to FM-UE capturing different types of abnormality in each group. In other words, in spite of similar overall FM-UE scores, could there be systematic differences in subcomponents of the FM-UE between subacute and chronic patients? We thus isolated and compared (a) the part of the FM-UE focusing on movement of the proximal arm (see Materials and Methods) and (b) the part of the FM-UE focusing on out-of-synergy movement between these two subgroups (subacute vs. chronic mild/moderate patients). We found no significant differences (proximal part: 30.8±1.0 vs. 29.7±1.6 for subacute vs. chronic, p=0.57; out-of-synergy part: 10.2±0.5 vs. 9.7±0.8 for subacute vs. chronic, p=0.60), ruling out that AMD^2^ differences could be explained by differences in the distribution of abnormality within the FM-UE for each group.

**Figure 5.**
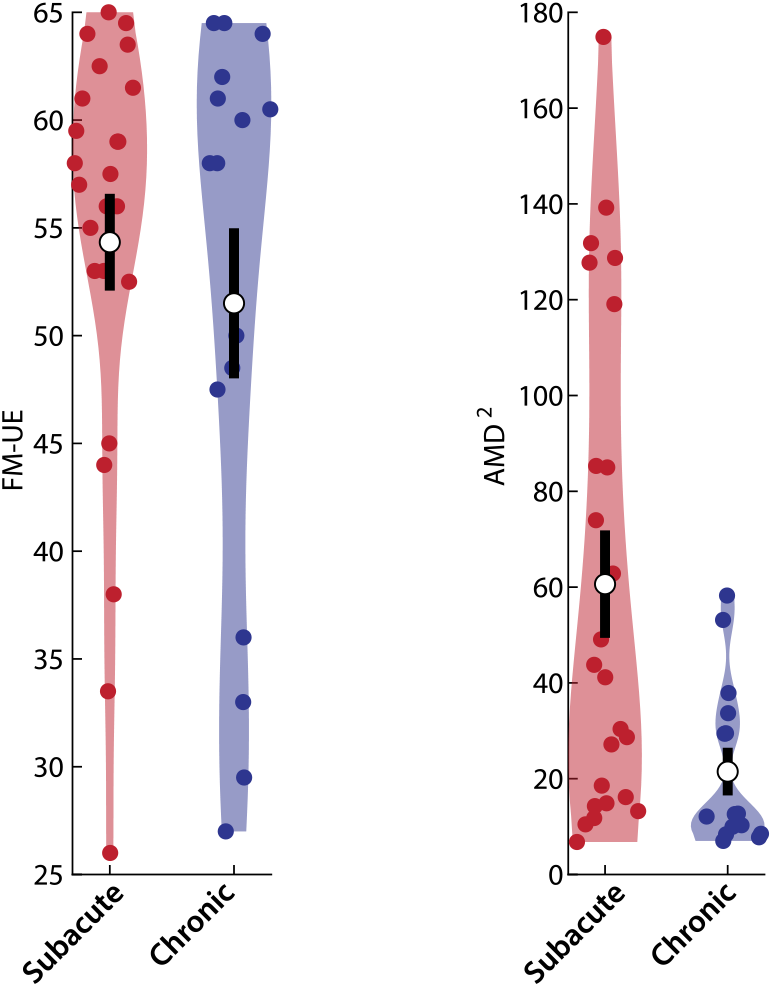
Subacute patients have worse kinematics compared to chronic patients in spite of matched Fugl-Meyer scores. Violin plots of the average FM-UE (left) and AMD^2^ (right) for chronic and subacute patients with moderate/mild (FM-UE≥26) impairment. Note how, for this FM range, where both subacute and chronic patients were adequately represented, we found much worse motor control (higher AMD^2^) for subacute patients compared chronic patients, despite matched FM-UE. White circles indicate mean values and thick black lines indicate mean ± SEM.

The difference in arm dexterity in spite of matched FM-UE that we observed, suggests that impaired arm dexterity (a negative sign) is dissociable from abnormal synergies (a positive sign on which FM-UE is based) – one is not causing the other. This failure on part of the FM-UE to capture differences in the quality of motor control during the subacute recovery stage further suggests that additional assessments may be needed to capture the full spectrum of the post-stroke motor control phenotype; and/or recovery of motor control within the subacute stage may lag behind corresponding improvements in FM-UE.

In addition to failing to capture motor control differences between the subacute and chronic groups, the FM-UE was a relatively poor predictor of kinematic deficits overall: while FM-UE accounted for a small fraction of the differences in AMD^2^ scores across the entire patient population (R^2^ = 0.27, p = 0.00007), this relationship appeared mostly driven by chronic patients with greater impairment (FM-UE<26) and large kinematic deficits (Figure 4B, blue dots towards the left; there was no significant relationship when only patients with mild/moderate impairment [FM-UE≥26] were examined, R^2^ = 0.04, p = 0.24). However, very low FM-UE scores could be attributable to damage that is extensive enough to separately lead to both an abnormal synergy and loss of dexterity, without meaning that loss of dexterity was specifically due to the abnormal synergy. Moreover, very low FM-UE scores imply substantial abnormality even for within-synergy movements, reducing the metric’s specificity as an indicator of the strength of abnormal synergies.

### As patients progressed through the subacute stage, the relationship between abnormal synergies and reaching dexterity increasingly resembled that of the chronic group

If the differences in kinematics between subacute and chronic patients matched for FM-UE were indeed due to time post-stroke, we would expect that, given time, the kinematics/FM-UE relationship for the subacute group would generally converge towards the kinematics/FM-UE relationship seen in the chronic group. To examine this, we tested for changes in both FM-UE and AMD^2^ in the subset of patients who completed the one-month follow-up (T2) session, and had been classified as moderate/high FMS (≥26) during the main (T1) session –14 patients in each group. We saw that, indeed, the relationship between AMD^2^ and FM-UE in the subacute group in the one-month follow-up (T2 session) tended to approach the relationship observed for chronic patients, with the most kinematically impaired subacute patients drastically reducing their AMD^2^ (illustrated by the long downward-facing red arrows in Figure 6A). On average, subacute patients improved both their FM-UE (59.6±1.1 vs. 63.2±0.7, p=0.0010) and their kinematics (AMD^2^ of 61.8±15.1 vs. 29.7±5.1, p=0.023). Changes in AMD^2^ between T1 and T2 might appear statistically weaker when tested using parametric tests due to lack of normality in the distribution of AMD^2^; a non-parametric comparison using bootstrap suggested a more clear difference (p = 0.0008). In contrast, chronic patients improved neither FM-UE nor kinematics (50.1±3.6 vs. 50.1±3.8, p = 1.00 for FM-UE and 23.4±4.6 vs. 21.2±4.7, p = 0.64 for AMD^2^ [p = 0.29 using bootstrap]), as illustrated in Figure 6B. We note that the lack of improvement in kinematics for the chronic group suggests no effect of savings or additional practice in the point-to-point reaching task (aside from a familiarization effect, illustrated in Figure S1), meaning that the changes in AMD^2^ we see in the subacute group represent improvements in reaching dexterity rather than motor learning.

**Figure 6.**
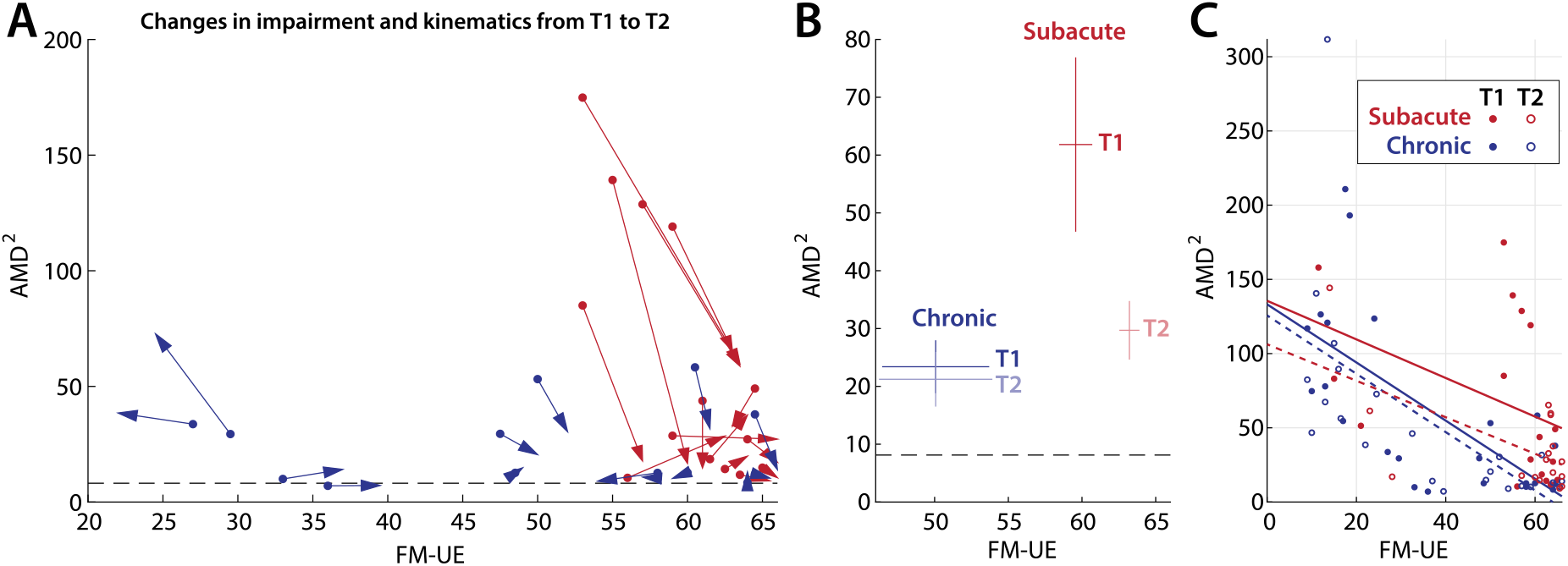
Changes in kinematics and FM-UE as recovery progresses. **A:** Individual changes in kinematic abnormalities (AMD^2^) and FM-UE between the main session (T1, dots) and the one-month follow-up (T2, tip of arrowpoints). **B:** Subject averages for these two groups. Errorbars indicate SEM. In both A and B, patients were only included if (a) they completed both T1 and T2 sessions, and (b) were classified in the moderate/mild group during T1. **C:** Linear fits of the FM-UE vs. AMD^2^ relationship for both subacute and chronic groups for both T1 and T2. In T2, the FM-UE / AMD^2^ for the subacute group becomes more similar to the chronic group.

In addition, we compared data from participants which completed both the T1 and T2 sessions regardless of FM-UE (a total of 17 subacute and 23 chronic) and investigated whether a stronger AMD^2^ /FM-UE relationship emerges among subacute patients in T2 compared to T1. Indeed, subacute participants showed a tighter FM vs. AMD^2^ relationship in T2 (R^2^=0.38, p = 0.0089) compared to T1 (R^2^ = 0.17, p = 0.10), more similar to the strength of the same relationship for chronic participants, which was strong in both T2 (R^2^=0.38, p = 0.0016) and T1 (R^2^=0.45, p = 0.00042), as shown in Figure 6C. This provides further evidence that the differences we observed between subacute and chronic groups in T1 were indeed due to time after stroke.

### Motor control deficits in the non-paretic arm were worse in the subacute compared to the chronic period

Finally, we analyzed trajectories in the non-paretic arm. While less pronounced than deficits in the paretic arm, both subacute and chronic patients had higher non-paretic AMD^2^ scores compared to controls (20.80±3.74 and 14.68±1.48 vs. 8.12±0.58 for the subacute, chronic, and controls, respectively, see Figure 7). An ANOVA revealed a significant effect of group (p = 0.0094). Post-hoc analyses showed that subacute patients had significantly greater AMD^2^ compared to controls (p = 0.0070) but not compared to chronic patients (p = 0.21). While chronic patients showed higher AMD^2^ than controls, this difference was not significant either (p = 0.25) –p-values obtained using the Tukey-Kramer test for multiple comparisons. This shows a clear deficit in motor control of the non-paretic arm at least early after stroke, mirroring previous results demonstrating reaching deficits in the non-paretic arm (Winstein and Pohl 1995; Haaland et al. 2004; Cortes et al. 2017). This finding suggests that loss of reaching dexterity after stroke is not restricted to the contralesional side, and may be captured by high-sensitivity assays like our simple reaching task and associated fPCA analysis.

**Figure 7.**
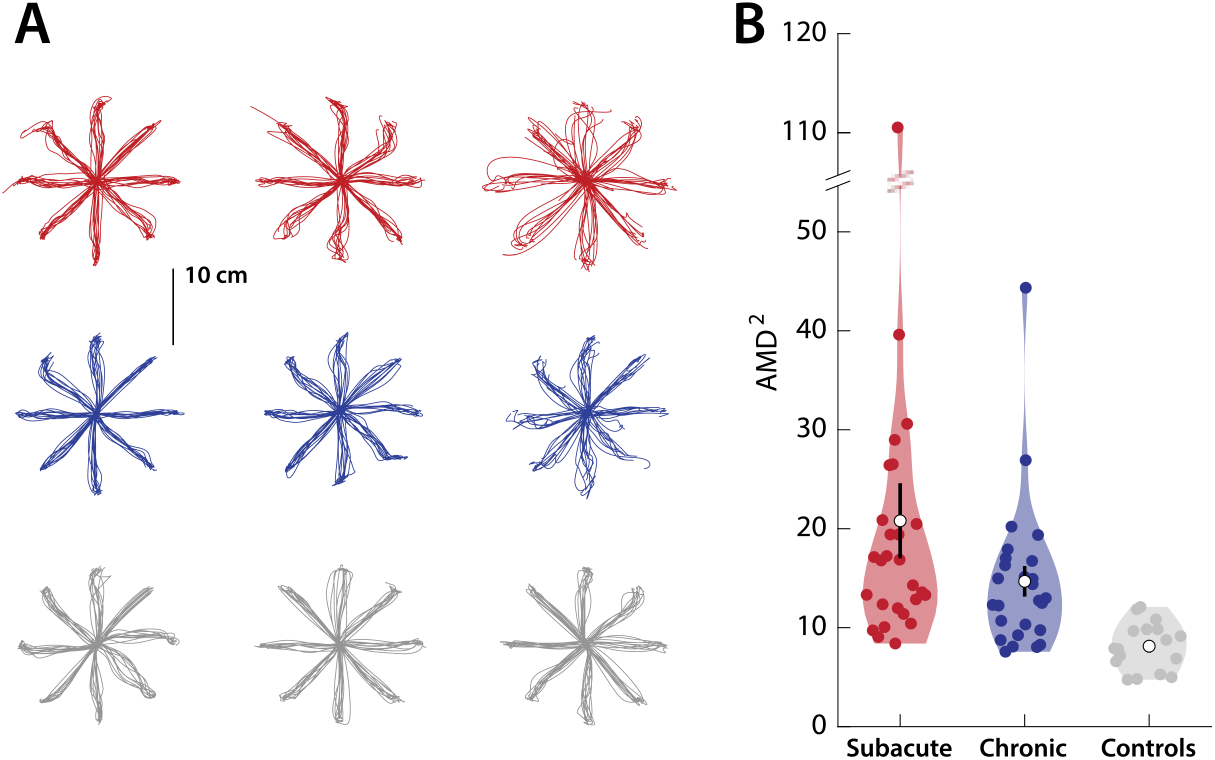
Kinematic abnormalities in the non-paretic arm. Subacute participants showed worse kinematics (higher AMD^2^) with their non-paretic arm compared to chronic participants and controls. **A:** example trajectories (same participants as in Figure 3B); **B:** Violin plots of the AMD^2^ for each group. White circles indicate mean values, thick black lines indicate mean ± SEM.

### Contributions of strength to the Fugl-Meyer score do not explain dissociation with reaching dexterity

Our findings suggest a dissociation between abnormal synergies, assessed through the FM-UE scale, and arm dexterity, assessed through kinematics. However, while FM-UE is a synergy-based measure, it may also reflect weakness – for example, we have previously found a strong correspondence between FM-UE improvement and recovery of strength (Cortes et al. 2017). Could the FM-UE vs. dexterity dissociation in our data instead reflect a strength/dexterity dissociation, as has been shown in earlier work (Ada et al. 1996)?

To assess the contribution of strength to our findings, we measured patients’ horizontal shoulder adduction/ abduction and elbow flexion/ extension strength. As dynamometry was only later added to the study assessments, it was not performed in all participants. In a subset of 17 participants (11 subacute and six chronic) plus an additional eight of our early participants (who were enrolled in a different study and were all at the chronic stage at the time of the measurement), we examined the relationship between FM-UE scores and strength at each joint (total of 25 participants). With the exception of horizontal shoulder adduction, strength correlated with FM-UE (see Figure 8). In particular, not only elbow extension strongly correlated with FM-UE (R^2^ = 0.68, p = 4×10^−7^), but elbow flexion also did (R^2^ = 0.39, p = 0.0009). The latter relationship was present even when we examined only the FM items strictly related to out-of-synergy movements (out-of-synergy FM-UE vs. elbow flexor strength: R^2^ = 0.35, p = 0.0019). As elbow flexor strength correlated almost as well as elbow extensor strength did to the part of FM-UE that evaluates movements *out* of flexor synergy, we find it unlikely that strength increases at the elbow are the cause of the ability to move out of synergy; rather, the correlations suggest a recovery process that is common for strength and synergies. We also found that elbow extensor strength correlated – even better than elbow flexor strength did – to the part of FM-UE that evaluates movements *within* flexor synergy (R^2^ = 0.51, p = 0.00007). Similarly, we find it unlikely that increased extension strength is the cause of the ability to move *within* flexor synergy; instead suggesting a common recovery process.

**Figure 8.**
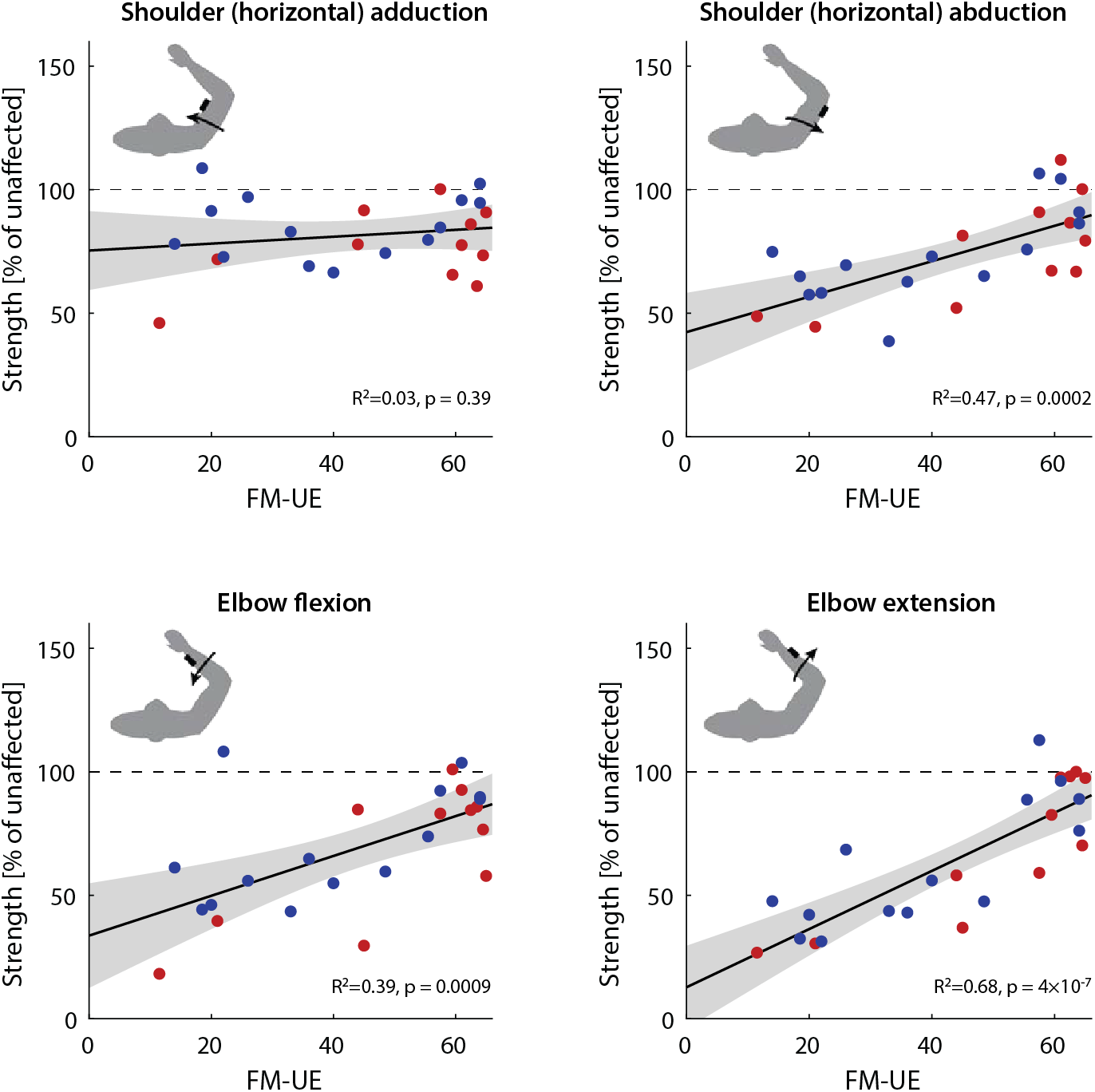
Relationship of weakness and FM-UE. Comparing strength (expressed as a % of non-paretic strength) with each patients’ FM-UE scores revealed strong correlations between FM-UE and shoulder abduction strength, elbow flexion strength, and elbow extension strength, but not shoulder adduction strength (which was relatively high across the whole patient population). For each panel, each dot represents a patient (red: subacute; blue: chronic, at the time of examination), whereas the solid black line indicates linear fit across all patients (with shaded area indicating the corresponding confidence interval).

These findings indicate that there is indeed a dissociation between synergy and arm dexterity rather than only a dissociation between strength and arm dexterity. Importantly, previous work has shown a strong correlation between FM-UE and measures of synergy abnormality measured through EMG (Bourbonnais et al. 1989; Dewald et al. 1995). This reinforces our conclusion that the findings here substantially reflect a dissociation between the presence of synergies and loss of dexterity. Future work may corroborate this using EMG, for example.

## Discussion

### Significance of our findings for assessing post-stroke motor impairment

Here we sought to dissect the hemiparesis phenotype into its constituent components. Given the prevalence and impact of hemiparesis, it is important to assess specific post-stroke motor impairments with quantitative metrics to enable clinicians to reliably characterize the initial deficit, and to predict and track recovery. This knowledge may help optimize rehabilitation (P. W. Duncan et al. 1992; C. Stinear 2010; C. M. Stinear et al. 2017) and also enable researchers to compare the effectiveness of different experimental treatments and interventions to enhance recovery. The FM is widely used in tracking post-stroke motor recovery, in fact it is the *de facto* impairment measure from the ICF used for studies and trials (Cortes et al. 2017; P. W. Duncan et al. 1992; Rabadi and Rabadi 2006; Chollet et al. 2011; Crisostomo et al. 1988; Francisco et al. 1998; Kwakkel et al. 2016; Van der Lee et al. 1999; Fasoli et al. 2003; Lo et al. 2010; Lum et al. 2002; P. Duncan et al. 2003; Feys Hilde M. et al. 1998). Here, however, we have shown that FM-UE might miss one of the components of hemiparesis - reaching dexterity loss – even though it might well capture weakness and abnormal synergies. Our findings, showing a poor correspondence between these two types of impairment, suggest the need for more careful matching between the rehabilitation intervention being tested in clinical studies and the chosen primary outcome measure: hemiparesis – or upper motor neuron syndrome – is too vague a term as it lumps weakness, synergies, and dexterity loss.

Why would FM-UE not be that suitable for assessing reaching dexterity deficits? Recent work has shown that reaching dexterity in stroke patients is improved with weight support; conversely, without weight support dexterity might be masked by weakness and abnormal synergies. Beer and colleagues (Beer et al. 2004) found that external arm support allowed for significantly greater peak torques when moving to distal targets requiring elbow extension and/or shoulder flexion, i.e. arm support facilitated movements that required breaking out of flexor synergy. At the same time, there was little, if any, effect of external arm support for movements to proximal targets, that involved elbow flexion and shoulder extension. A subsequent study by the same group also found that, while providing arm support allows for greater range and speed of elbow extension (Beer et al. 2007), this improvement was independent of reduced shoulder strength or elbow flexor/extensor strength imbalance. This suggested that abnormal synergies – and not merely weakness per se – were the sign alleviated by arm support. This finding mirrored earlier results for 3D movements (Zackowski et al. 2004), which showed a critical effect of synergy intrusion in the absence of weight support. Thus, tasks which require the patient to make multi-joint movements in 3D without support, like most of the test components of the FM-UE, will primarily reflect weakness and synergies, masking residual dexterity.

### Potential mechanisms behind differences in motor control between the subacute and chronic patients

Here, our data showed a clear dissociation between the FM-UE score and the quality of planar reaches. It still needs to be explained, however, why the dissociation took the *specific* form it did: it is not merely that FM scores are poor predictors of the quality of reaching, but there was a clear bias whereby planar reaches were substantially worse in the subacute as compared to the chronic group in patients with matched FM-UE scores. A potential explanation for this discrepancy may be that the residual corticospinal tract needs time and practice to reach its maximal level of potential performance. Thus, improvements in negative signs might lag improvements in positive ones. This explanation, however, appears to contradict our previous work showing that recovery of planar kinematics occurs over the first five weeks post-stroke and then plateaus (Cortes et al. 2017). We considered the potential explanation that at least some of the patients in the sub-acute group were still within this five-week window and therefore had not yet reached their full recovery. We reasoned that this could still be the case despite the fact that, overall, this group would be less impaired than the chronic group with respect to the FM-UE because they have matched scores and yet could only be expected to improve further. Indeed, the sub-acute patients with the highest AMD^2^ scores on T1 tended to improve drastically on T2, as illustrated in Figure 6A.

### The Fugl-Meyer assessment, abnormal synergies and weakness

In this study, we relied on FM-UE as a measure of abnormalities in muscle synergy. As we mention in the introduction, FM-UE was designed to capture the stages of post-stroke recovery described by (Twitchell 1951) and (Brunnstrom 1966), a prominent feature of which is the intrusion of abnormal synergies. In turn, this made the scale a strong indicator of synergy abnormality. In line with this idea, the degree of synergy abnormality assessed through EMG was found to strongly (negatively) correlate with FM-UE scores across patients (Bourbonnais et al. 1989; Dewald et al. 1995). Nevertheless, future studies could corroborate our findings with EMG.

In addition, here we also found that FM-UE scores also correlate with weakness, in line with our previous work suggesting that FM-UE recovery mirrors strength recovery (Cortes et al. 2017). This relationship was present not only for elbow extension strength but also for elbow flexion strength (Figure 8); it would be unlikely to have a causal relationship between increased flexion strength and ability to move out of flexor synergy. This observation is consistent with previous findings in which increased extensor reach was observed when arm support was given: externally-provided arm support is orthogonal to extensor (or flexor) strength (Sukal, Ellis, and Dewald 2007). Instead, ours and previous findings suggest that the FM-UE / weakness correlation might indicate a shared substrate between abnormal synergies and weakness.

The shared substrate hypothesis would fit with observations that abnormal synergies are mitigated when weakness is itself mitigated through arm support (Beer et al. 2004; 2007; Sukal, Ellis, and Dewald 2007). A potential explanation is that damage to the (contralateral) corticospinal tract (CST) after stroke may lead to increased reliance on the (ipsilateral) reticulospinal tract (RST), providing some strength at the expense of abnormal synergies (McPherson et al. 2018). In support of this theory, it was shown that corticospinal lesions in macaques led to increased responsivity in reticulospinal pathways which innervated forearm flexor muscles (Zaaimi et al. 2012); moreover it was recently shown that strength training in monkeys involves adaptations in the reticulospinal, but not the corticospinal tract (Glover and Baker 2020). This previous work, together with our findings, thus suggests an anatomical and physiological dissociation (corticospinal vs. reticulospinal) that may map onto the behavioral dissociation (positive vs. negative signs).

This theory, however, raises an apparent paradox: if upregulation of the RST increases both strength and abnormal synergies, then how do patients proceed to recover from synergies without concomitant loss of strength? There are a few possible non-mutually exclusive answers to this question that relate to recovery of CST function. First, as the CST becomes able to provide some strength, reliance on the RST may be reduced. Second, the RST might be unable to provide strength for some muscles, which instead may rely on CST recovery: for example, CST integrity may be necessary for strength in the FDI, a distal muscle but not the biceps (Schambra et al. 2019). Studies in monkeys using spike-triggered averaging have found that ipsilateral RST projections led to facilitation in the biceps but suppression in the triceps (Davidson and Buford 2004; Davidson, Schieber, and Buford 2007); thus, the (contralateral) CST – which can facilitate *either* flexors or extensors (Cheney, Fetz, and Mewes 1991)– may be necessary for control of extensor muscles, which is needed to break out of the flexor synergy. Third, given that both CST and RST converge upon spinal interneurons (Riddle, Edgley, and Baker 2009; Riddle and Baker 2010), the CST might directly regulate RST (Schepens and Drew 2006). Thus, CST recovery may restore its regulatory effect upon the RST, reducing abnormal synergies but maintaining strength.

### Limitations

Our comparison of kinematics between subacute and chronic groups focused on patients with moderate and mild impairment based on their FM-UE score. This was due to the very low number of subacute participants with low FM-UE scores, which prevented reliable comparisons with the corresponding part of the chronic group: we were able to recruit only 3 subacute patients with a FM-UE of <26 compared to 10 chronic patients in the same subgroup. We interpret this difference as the result of the difficulty of recruiting patients in the subacute stage after stroke. Both eligible subacute and chronic patients were recruited irrespective of their FM-UE score (within the 10-64 range). However, a subacute patient with high impairment will be likely to spend more time in the hospital and in rehabilitation, thus less likely to have time on research participation before the subacute time window expires. Moreover, increased impairment itself might make them less likely to be interested in joining in the first place, given that they may need more time to adjust. Hence, our observation in this study and others is that subacute patients who are able and eager to participate will tend to be less impaired in the first place.

However, we would not expect that the difference in kinematics between the subacute and chronic groups seen for moderate- and mild-impairment patients would also manifest in highly impaired patients. First, patients with high impairment would show difficulty with kinematics regardless of their recovery stage: lower and lower FM-UE scores would amount to impairment closer and closer to complete lack of movement. For example, a plegic patient would have a FM-UE score very close to zero, and being unable to move at all, they would have a complete deficit in their kinematics in our task, regardless of time after stroke. Second, the very fact that the FM-UE score failed to capture a good part of motor control deficits, specifically during the subacute stage, paired with the assumption that the subacute patients who participated may have had less *overall* impairment, suggests that subacute patients with low FM-UE may tend to fare better in components of impairment not well-captured by the FM-UE (one of which is motor control). Finally, there seems to be a difference how severe vs. moderate/mild patients recover: in contrast to moderate/mild patients, a fraction of severe patients may not recover much at all, and thus show less improvement as they advance to the chronic stage (Nakayama et al. 1994; Jørgensen et al. 1995; Krakauer and Carmichael 2017). While this observation was derived by comparing more general impairment scales (and FM-UE itself), it might also hold for movement kinematics as well.

We also note two more limitations. First, we did not formally examine patients’ sensory deficits, which could be a component of further study. Second, here we examined kinematics in 2D with full arm support. Fully evaluating post-stroke motor control, however, would need to examine kinematics in all three dimensions. 3D movement control may contain features not prominent in horizontal planar movement, such as the engagement of more muscles, a wider range of joint configurations, and dealing with the effects of weight bearing. For the latter part, especially, it may be worthwhile to systematically examine the relationship between FM-UE and kinematics under intermediate amounts of arm support rather than either full or none in order to properly map the interplay between synergies, strength, and kinematics.

## Conclusion

Here we show that there is a dissociation between loss of reaching dexterity and presence of abnormal muscle synergies in the contralesional arm after stroke; two prominent and characteristic signs of hemiparesis. To dissect these two signs, we designed a reaching task in which we isolated arm dexterity from synergy by providing weight support, and we separately assessed abnormal synergies using the Fugl-Meyer score for the Upper Extremity (FM-UE). Across patients, abnormal muscle synergies were not correlated with loss of arm dexterity. Critically, there was a large difference in dexterity deficits between subacute and chronic patients matched for similar levels of abnormal synergy: patients in the chronic stage had more normal planar reaching trajectories even with worse FM-UE scores. These dissociations suggest that abnormal synergies and dexterity deficits reflect distinct components of hemiparesis, perhaps attributable to damage to separable systems. Finally, we found that FM-UE scores correlated with arm weakness. This suggests that both synergies and weakness are independent of arm dexterity loss. In short, our findings suggest that recovery from hemiparesis does not proceed uniformly across its components. Stroke rehabilitation should be tailored for each patient based on their specific component deficits; a form of behavioral precision medicine.

## Data Availability

Data currently available upon request to the authors; will be uploaded to repository for easier access soon.

## Acknowledgements

This work has been supported by NIH (grant 5R01HD053793) and the Sheikh Khalifa Stroke Institute. We would like to thank Jeff Goldsmith, Jing Xu, Adrian Haith, and Juan Camilo Cortes for helpful discussions, Amanda Therrien for help with experiment software, as well as Kahori Kita and Kendra Cherry-Allen for help with assessments.

