## Supplementary Materials for "Positive and negative stroke signs revisited: dissociations between synergies, weakness, and impaired reaching dexterity"

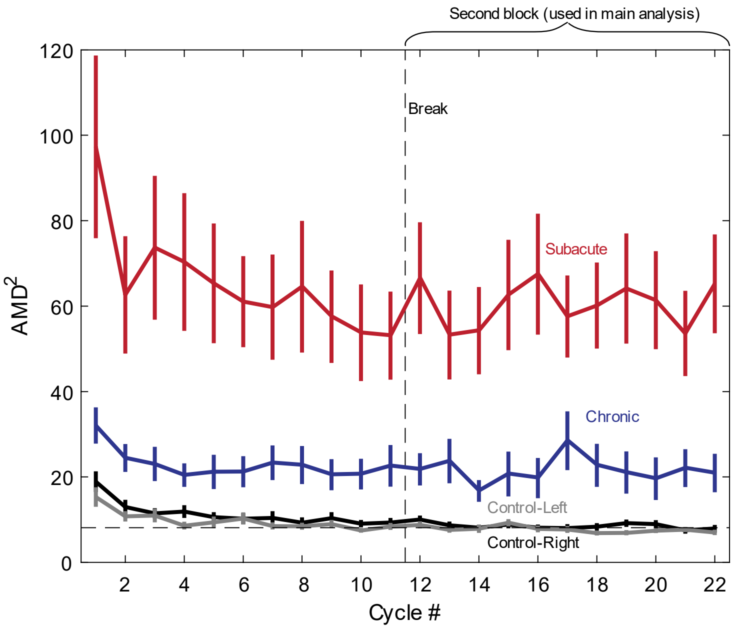


***Supplementary Figure 1. Cycle-by-cycle performance during the reaching task.*** *Shown are data from the paretic arms of subacute and chronic patients with moderate/mild (FM-UE>26) impairment, as well as from both arms of healthy controls. After 3-4 cycles, performance is stable. Note we used the second block (cycles 12-22) for our main analyses. The horizontal dashed line indicates baseline level (estimated as the average AMD² for controls during the second block).*

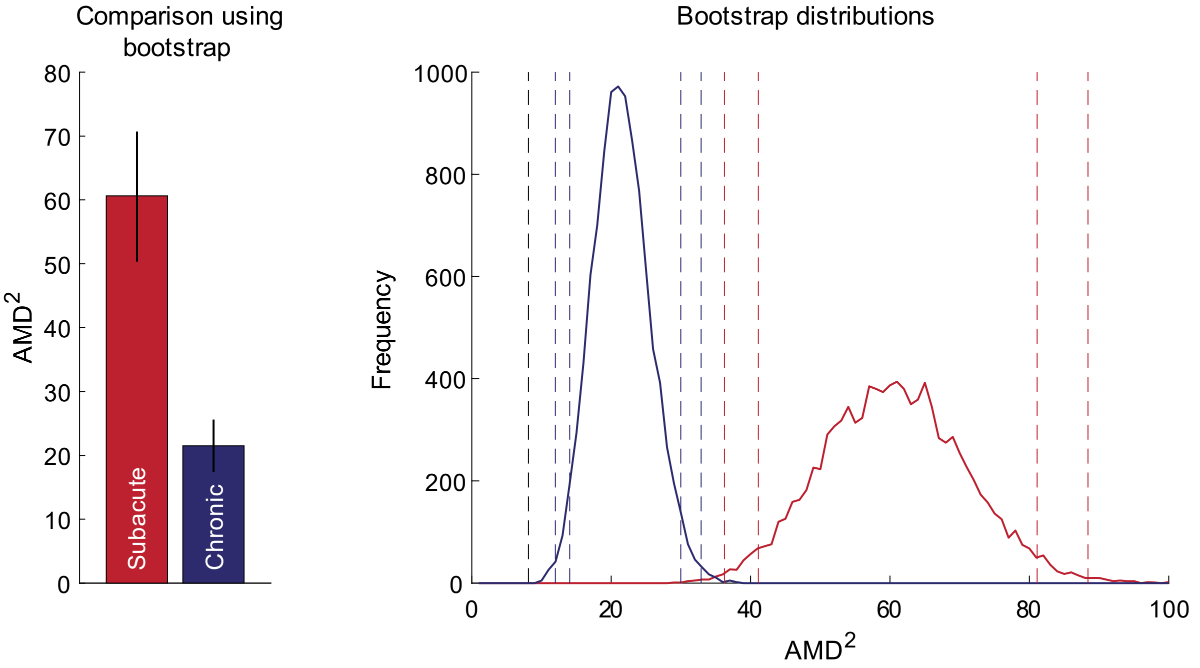


***Supplementary Figure 2. Comparison of AMD$^2$ in subacute vs. chronic stroke patients using bootstrap.*** *Instead of using a t-test as in the comparison illustrated in Figure 4, here we used a bootstrap procedure for the same data (paretic reaches of patients with moderate/mild (FM-UE>26) impairment).* ***Left:*** *Estimated mean±SEM [mean±standard deviation of bootstrapped estimates] for AMD$^2$; out of 10,000 bootstrap iterations, none showed higher average AMD$^2$ for chronic compared to subacute patients.* ***Right:*** *distribution of bootstrap results for the subacute (red) and chronic (blue) patients. Gray dashed line indicates baseline; colored dashed lines indicate the 0.5$^{th}$, 2.5$^{th}$, 97.5$^{th}$, and 99.5$^{th}$ percentiles of their corresponding distributions.*
